# Anti-thyroid drug use during the first trimester of pregnancy and the risk of birth defects in offspring: systematic review and meta-analysis of observational studies with methodological considerations

**DOI:** 10.1101/2020.06.10.20127233

**Authors:** Daniel R. Morales, Lionel Fonkwen, Hedvig M. Nordeng

## Abstract

**Background:** Maternal anti-thyroid drug (ATD) use during the first trimester of pregnancy has been associated with an increased risk of birth defects in offspring. Uncertainty remains on the size of this risk and how it compares to untreated hyperthyroidism due to methodological limitations of previous studies.

**Methods:** Systematic review of MEDLINE and EMBASE identifying observational studies examining ATD use during the first trimester of pregnancy and risk of birth defects. Data were extracted on study characteristics, adjusted effect estimates and comparator groups. Effect estimates were pooled using a random-effects generic inverse variance method of analysis and absolute risk calculated.

**Results:** Seven cohort studies and one case–control study (involving 6212322 pregnancies and 388976 birth defects) were identified. Compared to unexposed women without hyperthyroidism, the association between ATD first trimester use and birth defects in offspring was: adjusted risk ratio [aRR] 1.16 95% CI 1.08-1.25 for propylthyoruacil (PTU); aRR 1.28 95% CI 1.06-1.54 for methimazole/carbimazole (MMI/CMZ); aRR 1.51, 95% CI 1.16-1.97 for both MMI/CMZ and PTU; and aRR 1.15 95%CI 1.02-1.29 for untreated hyperthyroidism. The risk of major birth defects per 1000 live births was: 9.6 for PTU; 16.8 for MMI/CMZ; 30.6 for both MMI/CMZ and PTU; and 9.0 for untreated hyperthyroidism.

**Conclusions:** When appropriately analysed this risk of birth defects associated with ATD use in the first trimester of pregnancy is attenuated. Although still elevated, the risk of birth defects is smallest with PTU compared to use of MMI/CMZ and may be similar to that of untreated hyperthyroidism.

## Introduction

Hyperthyroidism affects between 0.1% to 0.4% of pregnancies and left untreated, may have detrimental maternal and foetal consequences including increasing the risk of preeclampsia, intrauterine growth restriction, preterm birth and maternal heart failure.[1–3] The most common cause of hyperthyroidism in young women is Grave’s disease, which results from stimulation of the thyroid by thyrotrophin receptor stimulating antibodies resulting in elevated free thyroxine (T4) and/or triiodothyronine (T3) and low thyroid-stimulating hormone (TSH), and typically requires treatment.[4] In contrast, gestational transient hyperthyroidism resulting from thyroid gland stimulation by the human chorionic gonadotrophin hormone often requires no treatment.

Clinical guidelines recommending treatment of hyperthyroidism during pregnancy parallels that of non-pregnant women, which is largely through the use of antithyroid drugs (ATDs). The two commonly prescribed drugs for the management of hyperthyroidism in pregnancy are methimazole/carbimazole (MMI/CMZ) and propylthyouracil (PTU) that are considered equally effective.[5] Both ATDs can cross the human placenta resulting in uncertainty about the risk of birth defects. PTU has been the preferred ATD during the first trimester of pregnancy due to the rare safety concerns surrounding the possible teratogenic effects of MMI/CMZ. The reported MMI/CMZ embryofetopathy include aplasia cutis congenita (absence of a portion of skin, often localised on the head), craniofacial malformations (choanal atresia; facial dysmorphism), defects of the abdominal wall and gastrointestinal tract (exomphalos, oesophageal atresia, omphalo-mesenteric duct anomaly), and ventricular septal defect.[6] Consequently, clinical guidelines [7,8] recommend switching from MMI/CMZ to PTU in unplanned pregnancies for the first trimester and then using MMI/CMZ thereafter due to concerns of PTU hepatotoxicity.[9] Consequently, the role of PTU during pregnancy has been re-examined follow reports of maternal hepatotoxicity associated with use of this ATD and of birth defects [9,10]. These safety concerns recently led to updated warnings from the European Medicines Agency (EMA) on the use of PTU and MMI/CMZ during pregnancy.[11,12]

Two meta-analyses were recently published reporting that use of MMI/CMZ and PTU during pregnancy were associated with an increased risk of birth defects.[13,14] These meta-analyses used the number of cases and the number of people in the sampled population from observational studies to calculate pooled effect estimates for MMI/CMZ, PTU and both MMI/CMZ and PTU exposure during pregnancy. By analysing the data as if they were trials, those estimates represent crude associations that are likely to be confounded.[15] Heterogeneity may also occur due to design differences of those studies and further information from other types of comparator or reference groups may aid decision making. No analysis of specific subgroups of malformations were performed. The aim of this study was to examine the association with use of MMI/CMZ or PTU during pregnancy on the risk of birth defects in offspring, through systematic review and meta-analysis of data using more appropriate methods to support causal inference.

## Method

A review of MEDLINE and EMBASE was conducted using a pre-specified search strategy to identify all observational studies (cohort, case–control, and sibling studies) assessing the risk of birth defects associated with use of PTU or MMI/CMZ during pregnancy, published on or before 16 September 2019. The search strategy is reported in the online supplementary material. Titles and abstracts were screened and full texts of relevant articles assessed for eligibility by two authors. Only English language publications and published data were included.

Approaches to the type of confounding adjustment and study design were described. Methodological quality and risk of bias were evaluated for each study using the ROBINS-I tool [16]. This tool is designed to assess the strengths and weaknesses of non-randomised studies on the effects of interventions in regards to their risk of bias. In this regard, the tool covers seven distinct domains through which bias could be introduced: confounding, selection or participants into the study, classification of interventions, deviations due to intended interventions, missing data, measurement of outcome measures and selection of the reported result. The systematic review was registered on the EU Register of Post-Authorisation Studies (EUPAS30990) and reported according to PRISMA (Preferred Reporting Items for Systematic Reviews).[17]

### Data extraction

We extracted data from eligible studies for the following characteristics: study design, study population, sample size, type of comparator or reference group reported, and the accuracy and completeness of information on confounders. For each comparison, crude and adjusted regression effect estimates (odds ratios, hazard ratios and rate ratios) were identified with corresponding 95% confidence intervals (CIs). The outcomes of interest were the risk of birth defects in children following MMI/CMZ exposure, PTU exposure and both MMI/CMZ and PTU exposure during the first trimester of pregnancy.

### Comparator groups and alternative study designs

Confounding by indication may occur when the underlying indication is associated with the outcomes being studied. Using different comparator or reference groups may help to circumvent or minimise this potential confounding.[18,19] To examine the effect of different comparator groups on such confounding, regression effect estimates for the following pre-specified comparator groups were sought: (1) maternal exposure during pregnancy vs. all unexposed women (referred to as the classical comparison), (2) maternal exposure during the pre-pregnancy period (“discontinuers”) vs. all unexposed women (this exposure may act as a negative control and should theoretically be non-causal), (3) maternal exposure during pregnancy vs. a non-medicated disease comparison group (“disease comparison group”, this restriction may reduce confounding by indication or severity); (4) siblings discordant for prenatal medication exposure (this study design accounts for all time-fixed within-family confounding); (5) paternal exposure during pregnancy vs. all unexposed women (this exposure may act as a negative control and should theoretically be non-causal); (6) a non-medicated disease comparison group vs. all unexposed women (testing for confounding by indication or the direct effect of hyperthyroidism).[18,19]

### Analysis

The characteristics of each study and method of confounding adjustment for those reporting regression effect estimates were first described. The number of cases and total population from each study were then used to replicate the meta-analytical approach used by Li et al. using a Mantel-Haenzel fixed-effect model.[15] Adjusted effect estimates from all identified publications were then extracted, then transformed on the natural log scale and pooled using the generic inverse variance method of analysis.[13] Random-effects models were generated for each type of exposure and comparator group separately. When more than one study used the same data source, the largest study was initially selected with sensitivity analysis substituting this with other studies form the same data source. Odds ratios from case–control studies and hazard ratios from cohort studies were combined because they closely approximate each other.[20,21] For reporting, pooled effect estimates are subsequently referred to as risk ratios (RR) throughout. Publication bias was assessed by testing for funnel-plot asymmetry using the Egger test. Analyses were conducted in Review Manager 5.3 (Copenhagen: The Nordic Cochrane Centre, The Cochrane Collaboration, 2014).

To estimate absolute risk, the birth defect rate in unexposed women per 1000 live births was calculated in unexposed women without hyperthyroidism. The pooled effect estimates from the meta-analysis were then used to calculate the absolute risk of birth defects that may have occurred per 1000 live births for each comparison group of interest. Absolute risk was calculated for each study and also as an overall summary estimate.

## Results

The systematic review included a total of eight studies reporting regression estimates and seven studies without regression estimates (figure 1). The characteristics of these studies are presented in table 1 and supplementary table S1.[22–36] Of the studies reporting adjusted effect estimates, seven were cohort studies (involving 6195342 pregnancies and 388,864 birth defects) [22–29] and one was a case–control study (involving 16,980 pregnancies and 112 birth defects).[23] Two of these studies were conducted using the same overlapping populations in Denmark.[22,25] Of the comparator groups reported, five studies reported adjusted effect estimates for the risk of birth defects associated with use of MMI/CMZ during pregnancy versus all unexposed women (the classical comparison), and one study compared MMI/CMZ use to a disease comparison group (supplementary table S2). The corresponding number of studies for PTU were five with a classical comparison group and two with a disease comparison group, respectively. For use of both MMI/CMZ and PTU during the first trimester of pregnancy (i.e. switching), three studies reported adjusted effect estimates for the classical comparison. No studies were identified that undertook a sibling study design, or examined pre-pregnancy exposure or paternal exposure. However, one study used a comparator consisting of either pre-pregnancy and post-pregnancy exposure.

**Figure 1.**
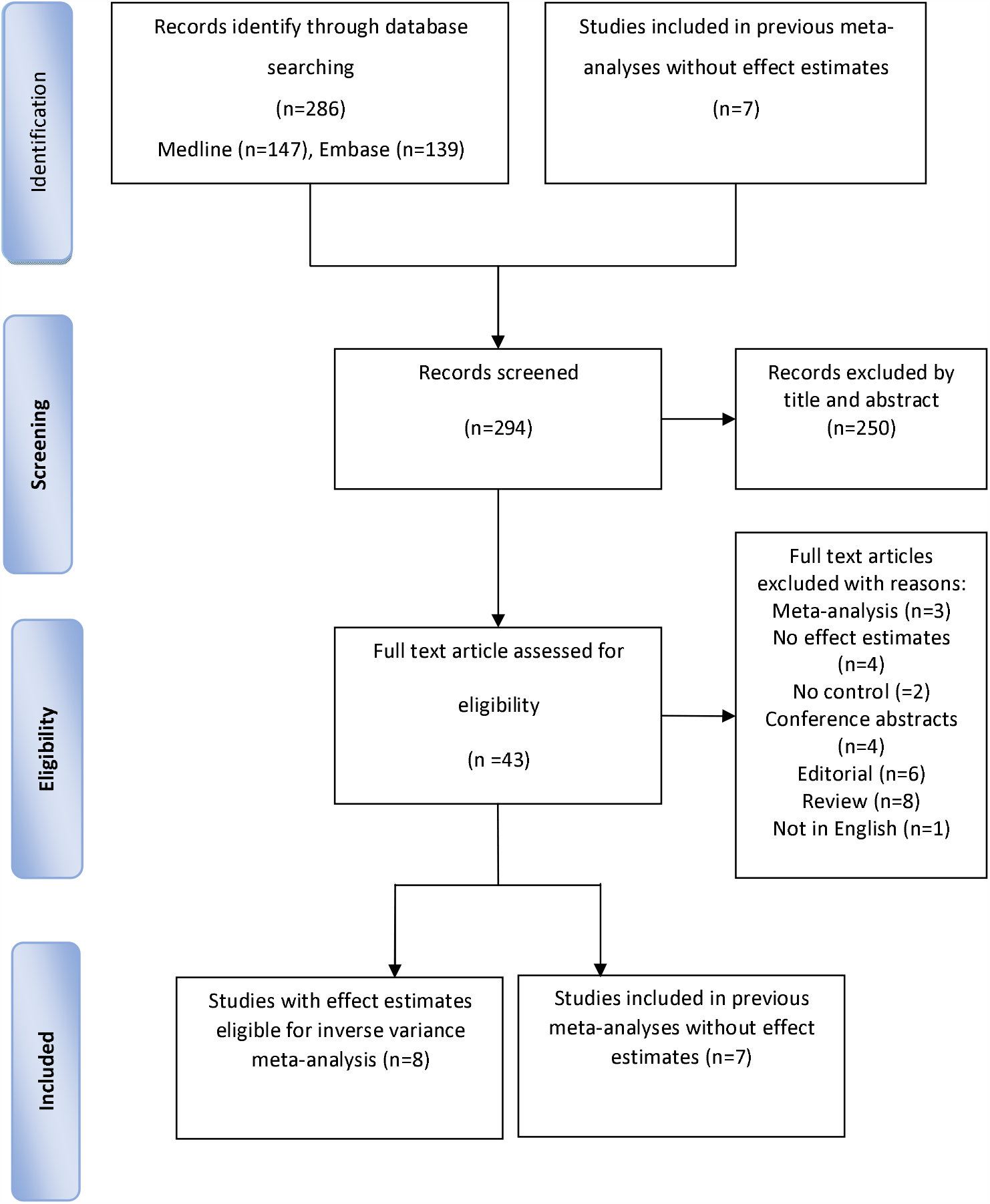
PRISMA (Preferred Reporting Items for Systematic Reviews) flow diagram for study selection process for the meta-analysis.

**Table 1.** Characteristics of the eight observational studies with effect estimates for risk of birth defects included in the meta-analysis.

| Author, publication year (Reference) | Type of study (Design and data source) | Country | Study population | Type of malformation** | Timing of exposure | Duration of follow-up in children | ATD exposure*** (cases/total) | Non-medicated women with hyperthyroidism (cases/total) | Non-medicated women reference group (cases/total) |
| --- | --- | --- | --- | --- | --- | --- | --- | --- | --- |
| Andersen 2019 <sup>20</sup> | Cohort study, National registries and birth cohorts | Denmark | Live births | All malformations and 8 malformation subgroups diagnosed before 2 years of age. | 6 months before pregnancy to gestational week 10 | Up to 2 years | PTU (74/889)<br>MMI (151/1574) | Unexposed to ATD (174/951) | Unexposed to ATD (77,791/ 1,159,181) |
| Seo 2018 <sup>21</sup> | Cohort study, Claims database | Korea | Live births | Major malformations and 10 birth defect subgroups | During the first trimester | Up to 1 year | PTU (699/9930)<br>MMI (91/1120)<br>MMI/PTU (147/1841) | - | Unexposed to ATD (170 716/ 2 872 109) |
| Andersen 2017 <sup>22</sup> | Cohort study, National registries | Sweden | Live births | All malformations and 13 malformation subgroups | 6 months before pregnancy to the end of gestational week 10 | Up to 2 years | PTU (14/218)<br>MMI (11/162)<br>MMI/PTU (4/66) | ATD prior pregnancy and after pregnancy (136/1551) | Unexposed to ATD (54827/682343) |
| Andersen 2013 <sup>23</sup> | Cohort study, National registries | Denmark | Live births and stillbirths | All malformations and 8 malformation subgroups | 6 months before pregnancy to the end of gestational week 10 | Up to 2 years | PTU (45/564)<br>MMI/CMZ (100/1097)<br>MMI/PTU (16/159) | ATD prior pregnancy and after pregnancy (190/3543) | Unexposed to ATD and without hyperthyroidism (45982/811730) |
| Korelitz 2013 <sup>24</sup> | Cohort study, Claims database | United states | Live births | Any malformation | 6 months before pregnancy to the end of pregnancy | Up to 1 year | PTU (66/915)<br>MMI (6/108)<br>MMI/PTU (14/126) | Thyrototoxicosis before/during/ after pregnancy (390/5932) | Unexposed to ATD and without hyperthyroidism (37351/634858) |
| Yoshihara 2012 <sup>25</sup> | Cohort study, Hospital database | Japan | Live births and perinatal losses of women with Graves' disease | Major malformations | During the first trimester | 1st visit post-delivery | PTU (26/1578)<br>MMI (50/1426) | Graves' disease (40/2065) | - |
| Chen 2011 <sup>26</sup> | Case-control Study, Claims database | Taiwan | Live births of women with hyperthyroidism and age matched controls without | Major malformations | During pregnancy | Not reported | PTU (5/630)<br>MMI (0/73) | Women with hyperthyroidism (15/2127) | Unexposed to ATD and euthyroid (92/14150) |
| Rosenfeld 2009 <sup>27</sup> | Prospective cohort study by Teratology Information Service | Israel | Live births and elective terminations*<br>Controls exposed to nonteratogens | Major malformations | Gestational weeks 4-13 | 0-6 years | PTU (1/80) | - | Exposed to non-teratogenic drugs (34/1066) |

ATD=Antithyroid drug. CMZ=carbimazole. MMI=methimazole. PTU=propylthiouracil. \* Elective terminations of pregnancy due to prenatally diagnosed anomalies. \*\*Malformations due to genetic disorders included. \*\*\*Andersen 2013: MMI/CMZ followed by PTU (n = 149) or PTU followed by MMI/CMZ (n = 10). Andersen 2017: MMI/CMZ followed by PTU (n=64) or PTU followed by MMI/CMZ (n=2). Korelitz 2013. Seo 2018: switching order not specified. Chen 2011: 516/630 and 65/73 received PTU and MMI in the first trimester respectively.

### Meta-analysis

The meta-analytical approach used by Li et al. was first replicated using the number of reported cases and total population with a Mantel-Haenzel fixed-effect model. When analysed in this way, maternal MMI/CMZ exposure and PTU exposure during the first trimester of pregnancy was associated with a significantly increased risk of birth defects in offspring compared to the classical reference group of all unexposed women (RR 1.20 95%CI 1.02-1.42 for PTU; RR 1.64 95%CI 1.39-1.92 for MMI/CMZ; RR 1.83 95%CI 1.30-2.56 for MMI/CMZ and PTU use, supplementary figure S1).

When effect estimates that adjusted for confounders were pooled using the inverse variance method of analysis, maternal use of MMI/CMZ during the first trimester of pregnancy was associated with a smaller but still significantly increased risk of birth defects compared to all unexposed women (random-effects adjusted risk ratio [RaRR] 1.28 95% CI 1.06-1.54, figure 2). This was similarly the case for use of PTU (RaRR 1.16 95% CI 1.08-1.25) and exposure to both MMI/CMZ and PTU during the first trimester of pregnancy (RaRR 1.51 95% CI 1.16-1.97).

**Figure 2.**
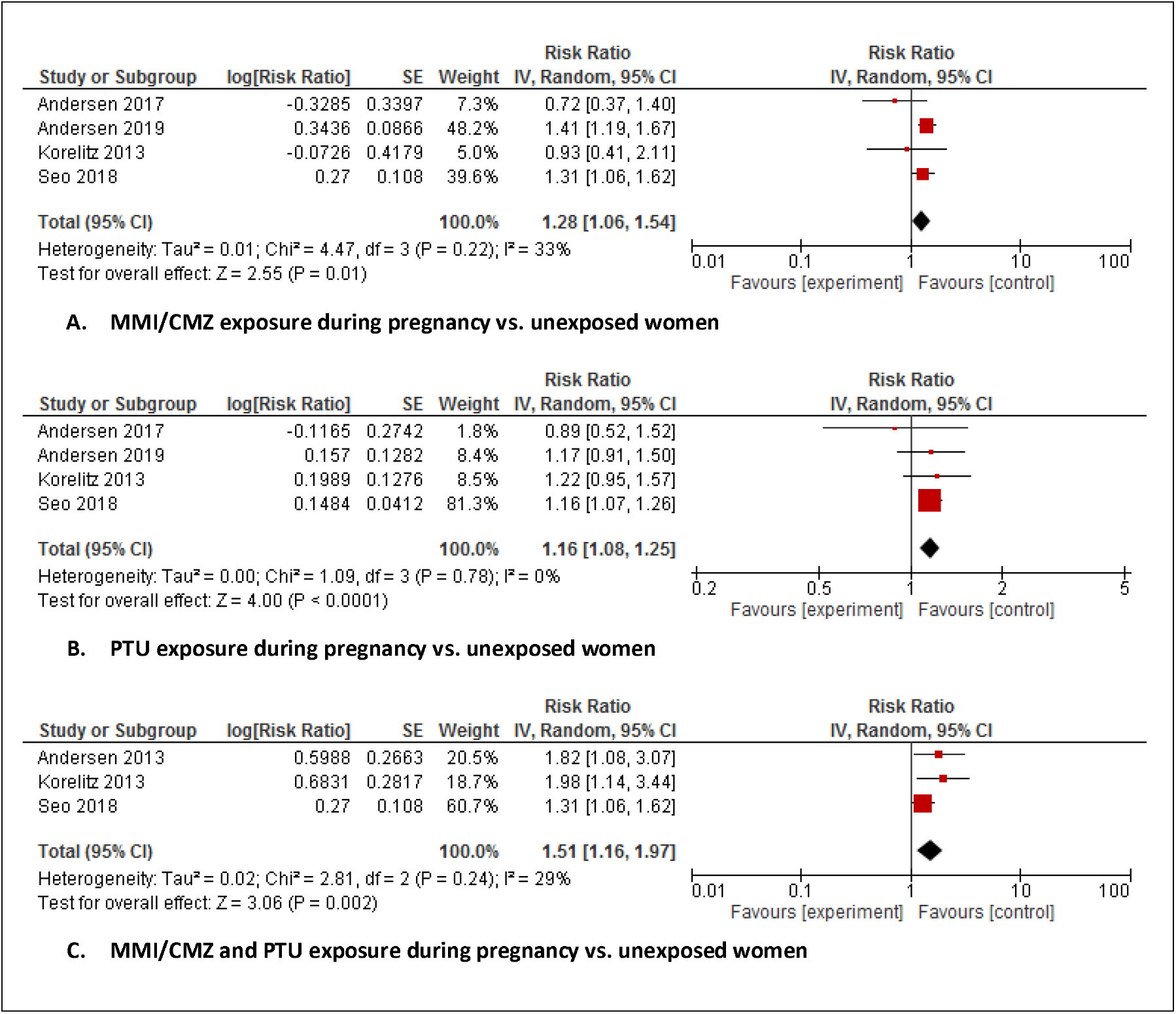
Association between maternal ATD exposure during the first trimester of pregnancy and risk of congenital anomalies in offspring compared to untreated women without hyperthyroidism when analysing adjusted effect estimates.

The risk of birth defects among unexposed women with hyperthyroidism compared to unexposed women without hyperthyroidism was also significantly elevated (RaRR 1.15 95%CI 1.02-1.29, figure 3). When PTU exposure was compared to unexposed women with hyperthyroidism reported in two studies, PTU exposure during the first trimester of pregnancy was not associated with an increased risk of birth defects (RaRR 0.72 95% CI 0.47-1.11, figure 3). In contrast, only one study reported this comparison for MMI/CMZ that was significantly elevated (RR 2.28, 95% CI 1.54 - 3.33).

**Figure 3.**
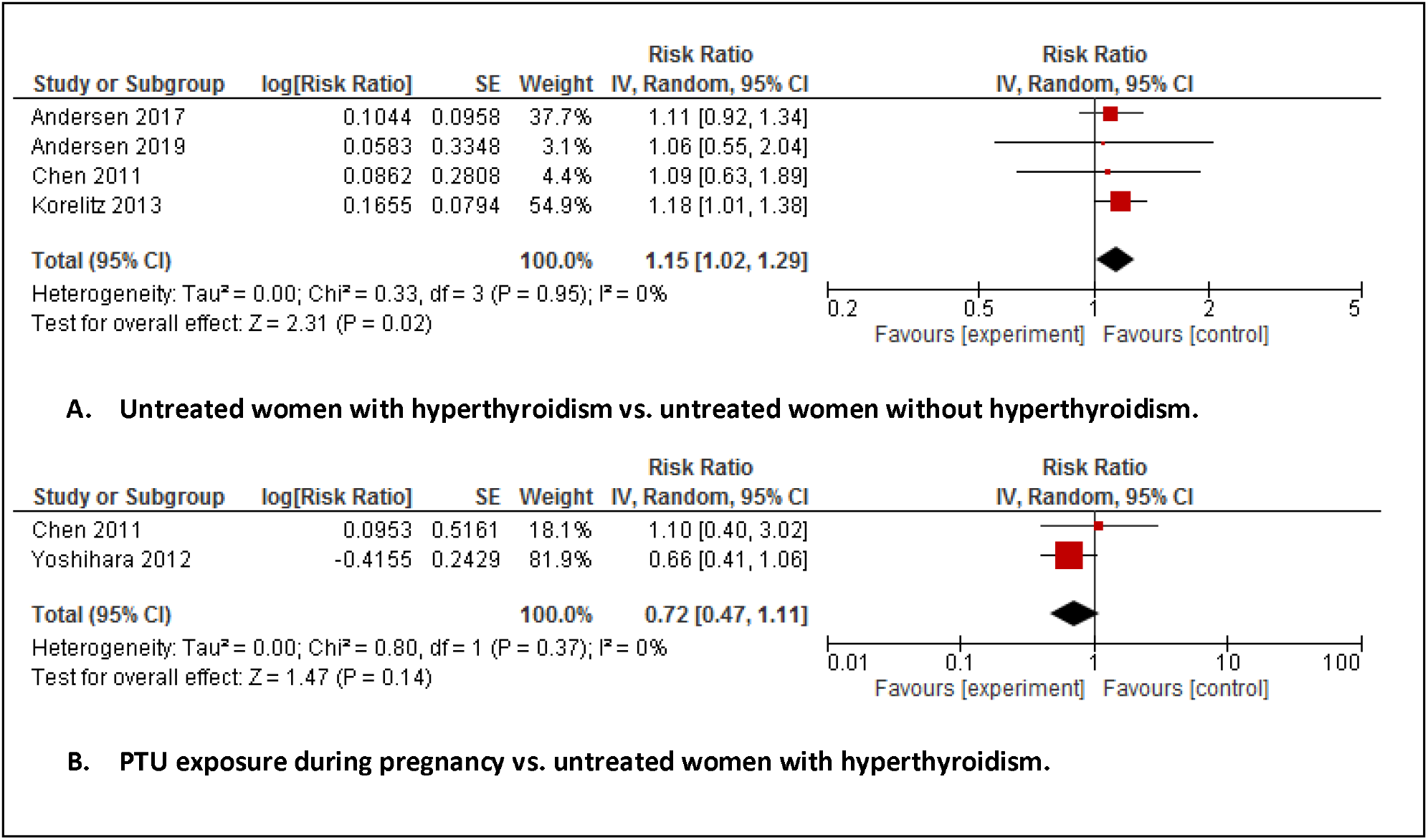
Associations between the risk of congenital anomalies in offspring of non-medicated women with hyperthyroidism compared to all non-medicated women without hyperthyroidism (A) and PTU exposure during the first trimester of pregnancy compared to all non-medicated women with hyperthyroidism (B).

### Sensitivity analyses

When studies conducted using the same data source were substituted, pooled effect estimates for MMI/CMZ exposure and PTU exposure were RaRR 1.28 (95CI 0.96-1.71) and RaRR 1.17 (95%CI 1.09-1.26) respectively. When studies reporting effect estimates for subgroups of birth defects were substituted [24], pooled effect estimates for MMI/CMZ and PTU groups were RaRR 1.36 (95%CI 1.05-1.76) and RaRR 1.17 (95%CI 1.09-1.26) respectively.

### Absolute risk

The overall rate of birth defects in unexposed women without hyperthyroidism was 59.9 (95%CI 59.7-60.1) per 1000 live births (table 2). The absolute risk difference in the number of birth defects associated with each comparison group was: 9.0 in women with non-medicated hyperthyroidism; 9.6 in women treated with PTU; 16.8 in women treated with MMI/CMZ; and 30.6 in women treated with both MMI/CMZ and PTU during the first trimester of pregnancy (table 2).

**Table 2.** Absolute risk of birth defects per 1000 live births associated with MMI/CMZ exposure, PTU exposure and non-medicated hyperthyroidism during pregnancy.

| Country (reference) | Total number of pregnancies | Major malformation rate in unexposed women (per 1000 live births) | Estimated excess number of malformations (per 1000 live births) |  |  |  |
| --- | --- | --- | --- | --- | --- | --- |
|  |  |  | Non-medicated hyperthyroidism | PTU exposure | MMI/CMZ exposure | MMI/CMZ and PTU exposure |
| Denmark<sup>20</sup> | 1 243 353 | 67.1 | 10.1 | 10.7 | 18.8 | 34.2 |
| Korea<sup>21</sup> | 2 886 970 | 59.4 | 8.9 | 9.5 | 16.8 | 30.3 |
| Sweden<sup>22</sup> | 684 340 | 80.4 | 12.1 | 12.9 | 22.5 | 41.0 |
| Denmark<sup>23</sup> | 817 093 | 56.7 | 8.5 | 9.1 | 15.9 | 28.9 |
| United states<sup>24</sup> | 904 497 | 58.8 | 8.8 | 9.4 | 16.5 | 30.0 |
| Taiwan<sup>26</sup> | 16 980 | 6.5 | 1.0 | 1.0 | 1.8 | 3.3 |
| Israel<sup>27</sup> | 1 256 | 31.9 | 4.8 | 5.1 | 8.9 | 16.3 |
| Overall* | 6 561 233 | 59.9 | 9.0 | 9.6 | 16.8 | 30.6 |

\*Overall rate calculated excluding Andersen 2013 due to overlapping population.
Yoshihara<sup>25</sup> excluded as no data on malformation rate in unexposed women without hyperthyroidism.

### Assessment of confounding factors and risk of bias

Of the eight studies, six adjusted for maternal age, and three for each of the following: the infant’s sex; year of birth; parity; and pregnancy type (supplementary table S3). Adjustment for maternal physical history and smoking status occurred in three and two studies respectively. Adjustment of maternal physical history consisted mainly of metabolic conditions including diabetes and hyperlipidaemia. No studies adjusted for alcohol use during the first trimester of pregnancy and only one study adjusted for any other type of medication use during the first trimester of pregnancy, namely antiepileptic use.

For the classical comparison using all unexposed women, the risk of bias varied according to the domain studied and was influenced by variation in the type of confounding adjustment undertaken and uncertainty around exposure and outcome ascertainment bias (supplementary table S4). The Egger test for MMI/CMZ exposed vs. unexposed (p value = 0.020), PTU exposed vs. unexposed (p-value= 0.005) and PTU and MMI/CMZ exposed vs. unexposed (p-value = 0.027) were statistically significant for funnel plot asymmetry although the number of included studies evaluating this were limited.

## Discussion

Meta-analysis is a powerful tool that combines estimates from multiple studies to improve power, precision, and answer questions that are limited by individual studies. By meta-analysing data from observational studies of ATD therapy during pregnancy as if they were trials clearly overestimates risk, which attenuates when more appropriate methods are used.[13] Furthermore, although observed associations between birth defects and exposure to PTU and/or MMI/CMZ during the first trimester of pregnancy were still significantly elevated, associations between women using PTU and those with non-medicated hyperthyroidism were similar in size, whilst the highest estimates were observed in the groups switching between MMI/CMZ and PTU.

PTU is now typically considered the safer option when hyperthyroidism requires treatment during the first trimester of pregnancy. However, safety concerns surrounding PTU exposure and the risk of hepatic toxicity and birth defects have emerged leading to regulatory updates to the product information in 2011 and 2019, respectively.[10,12] Our findings for PTU exposure during the first trimester of pregnancy are reassuring as it suggests that PTU exposure does not increase the risk of birth defects beyond that of the underlying untreated maternal disease, but potentially has the additional advantage of reducing maternal morbidity associated with non-medicated hyperthyroidism according to previous research.[5] We estimated that the excess number of major birth defects with use of PTU was 9.6 per 1000 live births compared with 9 in non-medicated women with hyperthyroidism.

The highest risk of birth defects was associated with use of both MMI/CMZ and PTU during the first trimester of pregnancy. This corresponds to women who are switched from one product to the other. The effects of switching can be difficult to interpret with these data as the order of switching was not always specified although in the majority of instances switches appeared to be appropriate as most switching was from MMI/CMZ to PTU during the first trimester of pregnancy. Given current guideline recommendations, this could represent a group more likely to include women with unplanned pregnancies that can differ from pregnancies that are planned in terms of lifestyle and early folic acid use for example.

Our analysis clearly demonstrates that only by using adjusted effect estimates can information from measured confounding factors be appropriately accounted for and that risk estimates from meta-analyses based upon crude numbers may be of limited value for clinical and regulatory decision making.[14,15] We recommend avoiding this method of meta-analysis for non-randomised studies when adjusted effect estimates from observational studies are available. This is particularly important when such estimates are used to calculate absolute risk. In this regard, individual observational studies should be properly designed and conducted to provide the appropriate primary data, including being adequately powered to reduce the risk of a type-II error.

This study has several potential limitations. First, although accounting for confounders attenuated the observed associations, the included studies were heterogeneous in their approach to confounding adjustment suggesting that residual confounding remains possible. Other features that affected the risk estimates include the lack of exclusion of offspring with birth defects due to genetic disorders. This may introduce bias if women with hyperthyroidism/Graves disease have an increased risk of having children with birth defects disorders of genetic origin.[37] Moreover, ascertainment bias related to the outcome is expected as clinicians may scrutinize children of women with hyperthyroidism to a greater extent than children of health women, especially after the signals of ATD teratogenicity. If this were to happen this could have a larger effect on studies with less follow up to detect outcomes of interest although we observed no obvious impact of this with the included studies. For these reasons it is of great importance to use a non-exposed disease comparison group as an alternative comparator to better infer causality.[18,19] Notably no sibling study designs have been reported despite the potential advantage of controlling for shared familial confounding by design.[19] Moreover, the majority of studies assessed only malformations in live borns and therefore miss birth defects due to elective termination, miscarriages or stillbirths. Conditioning on live births may give rise to collider bias if both exposure and outcome are associated with survival.[38] Second, despite the advantages of having a non-medicated disease comparison group, this approach may not account for differences in the severity or type of hyperthyroidism. It is likely that transient gestational hyperthyroidism may represent a significant proportion of untreated cases whereas Grave’s disease will most likely be medicated. Third, despite demonstrating comparable risk from PTU exposure and non-medicated hyperthyroidism, the overall number of studies was small and further observational studies may be useful, including those that explore the relationships between different comparator groups and study designs. We also cannot exclude the possibility of publication bias. Fifth, this study examines birth defects as a composite outcome and data may not be generalizable to subtypes of birth defects, and especially MMI embryofetopathy. However, whilst MMI/CMZ may be associated with a larger risk of birth defects not all studies consistently reported an elevated risk demonstrating that some uncertainty about this association remains. For example, in a large study from Swedish there were no cases of aplasia cutis and choanal or esophageal atresia that have described as being the MMI embryopathy.[15] The only study that was sufficiently powered to assess this malformation pattern however, found 5 additional cases of MMI embroypathy per 10 000 after first trimester exposures to ATDs compared to non-exposed (0.12% vs. 0.07%).[23] Only one study reported a dose-response relationship which observed larger risk estimates for congenital anomalies with high cumulative doses of CBZ/MMI but not with PTU.[9] However, this study was not sufficiently powered to examine a dose-response relationship adequately and further evidence is still required.

Assuming MMI/CMZ and PTU are equally effective and no significant unmeasured confounding, our findings suggest that PTU use is the safer option in terms of the risk of birth defects in offspring, whilst non-medicated hyperthyroidism may also carry a risk. This suggests that the net-benefit of managing hyperthyroidism during the first trimester of pregnancy may be in favour of using PTU when clinically indicated. However, such use must also be balanced against other potential risks with PTU such as hepatoxicity and the limitation of current observational studies.

## Conclusion

Previous meta-analyses examining the risk of maternal ATD use during the first trimester of pregnancy overestimate the risk of birth defects in offspring. When appropriately analysed this risk is attenuated and although still elevated the risk of birth defects is smallest associated with use of PTU during the first trimester of pregnancy.

## Data Availability

All data is either contained within the manuscript or can be accessed via the referenced publications.

## Author contributions

DM and LF collected the data and performed the analysis with guidance from HN. DM is the guarantor for the study. All authors contributed to the study design, interpretation of results, writing the manuscript and approved the final draft.

## Disclaimer

This work has been deposited as a preprint. It has not been peer reviewed and the work, its content and/or its conclusions may be subject to change. Caution is therefore advised over its use in decision making. The views expressed in this article are the current personal views of the authors and may not be understood or quoted as reflecting the views of any organisation or one of its committees or working parties.

## Study registration

The study has been registered in the EU PAS Register (no. EUPAS30990) [www.encepp.eu)

## Ethical approval

No ethical approval was required to conduct this study.

## References

1. Davis LE, Lucas MJ, Hankins GD, Roark ML, Cunningham FG. Thyrotoxicosis complicating pregnancy. Am J Obstet Gynecol. 1989 Jan;160(1):63–70.

2. Millar LK, Wing DA, Leung AS, Koonings PP, Montoro MN, Mestman JH. Low birth weight and preeclampsia in pregnancies complicated by hyperthyroidism. Obstet Gynecol. 1994 Dec;84(6):946–9.

3. Luewan S, Chakkabut P, Tongsong T. Outcomes of pregnancy complicated with hyperthyroidism: a cohort study. Arch Gynecol Obstet. 2011 Feb;283(2):243–7.

4. Lazarus JH. Hyperthyroidism during pregnancy: etiology, diagnosis and management. Women’s Health 2005;1:97–104.

5. Alexander EK, Pearce EN, Brent GA, Brown RS, Chen H, Dosiou C, Grobman WA, Laurberg P, Lazarus JH, Mandel SJ, Peeters RP, Sullivan S. 2017 Guidelines of the American Thyroid Association for the Diagnosis and Management of Thyroid Disease During Pregnancy and the Postpartum. Thyroid. 2017 Mar;27(3):315–389.

6. Methimazole embryofetopathy. The portal for rare diseases and orphan drugs. Available at: https://www.orpha.net/consor/cgi-bin/OC_Exp.php?Lng=GB&Expert=1923 Accessed 04/12/2019.

7. Bahn RS, Burch HS, Cooper DS, Garber JR, Greenlee CM, Klein IL, et al. The role of propylthiouracil in the management of Graves’ disease in adults: report of a meeting jointly sponsored by the American Thyroid Association and the Food and Drug Administration. Thyroid. 2009;19:673–4.

8. De Groot L, Abalovich M, Alexander EK, Amino N, Barbour L, Cobin RH, et al. Management of thyroid dysfunction during pregnancy and postpartum: an Endocrine Society clinical practice guideline. J Clin Endocrinol Metab. 2012;97:2543–65.

9. Seo GH, Kim TH, Chung JH. Antithyroid Drugs and Congenital Malformations: A Nationwide Korean Cohort Study. Ann Intern Med. 2018;168(6):405–13.

10. U.S. Food and Drug Administration. FDA Drug Safety Communication: New Boxed Warning on severe liver injury with propylthiouracil. 2010. Accessed at https://www.fda.gov/drugs/postmarket-drug-safety-information-patients-and-providers/fda-drug-safety-communication-new-boxed-warning-severe-liver-injury-propylthiouracilon11/10/2019.

11. PRAC recommendations on signals. November 2018. Available at: https://www.ema.europa.eu/en/documents/prac-recommendation/prac-recommendations-signals-adopted-26-29-november-2018-prac-meeting_en.pdf. Accessed [01/10/2019].

12. PRAC recommendations on signals. June 2019. Available at: https://www.ema.europa.eu/en/documents/prac-recommendation/prac-recommendations-signals-adopted-11-14-june-2019-prac-meeting_en.pdf. Accessed [01/10/2019].

13. Reeves BC, Deeks JJ, Higgins JPT, Shea B, Tugwell P, Wells GA. Chapter 24: Including non-randomized studies on intervention effects. In: Higgins JPT, Thomas J, Chandler J, Cumpston M, Li T, Page MJ, Welch VA (editors). Cochrane Handbook for Systematic Reviews of Interventions version 6.0 (updated July 2019). Cochrane, 2019. Available from www.training.cochrane.org/handbook.

14. Song R, Lin H, Chen Y, Zhang X, Feng W. Effects of methimazole and propylthiouracil exposure during pregnancy on the risk of neonatal congenital malformations: A meta-analysis. PLoS One. 2017 Jul 3;12(7):e0180108. doi:10.1371/journal.

15. Li H, Zheng J, Luo J, Zeng R, Feng N, Zhu N, Feng Q. Congenital anomalies in children exposed to antithyroid drugs in-utero: a meta-analysis of cohort studies. PLoS One. 2015 May 14;10(5):e0126610.

16. Sterne JA, Hernán MA, Reeves BC, Savović J, Berkman ND, Viswanathan M, Henry D, Altman DG, Ansari MT, Boutron I, Carpenter JR, Chan AW, Churchill R, Deeks JJ, Hróbjartsson A, Kirkham J, Jüni P, Loke YK, Pigott TD, Ramsay CR, Regidor D, Rothstein HR, Sandhu L, Santaguida PL, Schünemann HJ, Shea B, Shrier I, Tugwell P, Turner L, Valentine JC, Waddington H, Waters E, Wells GA, Whiting PF, Higgins JP. ROBINS-I: a tool for assessing risk of bias in non-randomised studies of interventions. BMJ. 2016 Oct 12;355:i4919.

17. Moher D, Liberati A, Tetzlaff J, Altman DG, PRISMA Group. Preferred reporting items for systematic reviews and meta-analyses: the PRISMA statement. PLoS Med. 2009;6(7):e1000097.

18. Morales DR, Slattery J, Evans S, Kurz X. Antidepressant use during pregnancy and risk of autism spectrum disorder and attention deficit hyperactivity disorder: systematic review of observational studies and ethodological considerations. BMC Med. 2018 Jan 15;16(1):6.

19. Wood ME, Lapane KL, van Gelder MMHJ, Rai D, Nordeng HME. Making fair comparisons in pregnancy medication safety studies: An overview of advanced methods for confounding control. Pharmacoepidemiol Drug Saf. 2018 Feb;27(2):140–147.

20. D’Agostino RB, Lee ML, Belanger AJ, Cupples LA, Anderson K, Kannel WB. Relation of pooled logistic regression to time dependent Cox regression analysis: the Framingham Heart Study. Stat Med. 1990;9(12):1501–15.

21. Lanza A, Ravaud P, Riveros C, Dechartres A. Comparison of Estimates between Cohort and CaseControl Studies in Meta-Analyses of Therapeutic Interventions: A Meta-Epidemiological Study. PLoS One. 2016 May 9;11(5):e0154877.

22. Andersen SL, Knøsgaard L, Olsen J, Vestergaard P, Andersen S. Maternal thyroid function, use of antithyroid drugs in early pregnancy and birth defects. J Clin Endocrinol Metab. 2019 Aug 13. pii: jc.2019-01343.

23. Seo GH, Kim TH, Chung JH. Antithyroid Drugs and Congenital Malformations: A Nationwide Korean Cohort Study. Ann Intern Med. 2018 Mar 20;168(6):405–413.

24. Andersen SL, Olsen J, Wu CS, Laurberg P. Birth defects after early pregnancy use of antithyroid drugs: a Danish nationwide study. J Clin Endocrinol Metab. 2013;98(11):4373–81.

25. Andersen SL, Lonn S, Vestergaard P, Torring O. Birth defects after use of antithyroid drugs in early pregnancy: a Swedish nationwide study. Eur J Endocrinol. 2017;177(4):369–78.

26. Korelitz JJ, McNally DL, Masters MN, Li SX, Xu Y, Rivkees SA. Prevalence of thyrotoxicosis, antithyroid medication use, and complications among pregnant women in the United States. Thyroid : official journal of the American Thyroid Association. 2013;23(6):758–65.

27. Yoshihara A, Noh J, Yamaguchi T, Ohye H, Sato S, Sekiya K, et al. Treatment of graves’ disease with antithyroid drugs in the first trimester of pregnancy and the prevalence of congenital malformation. J Clin Endocrinol Metab. 2012;97(7):2396–403.

28. Chen CH, Xirasagar S, Lin CC, Wang LH, Kou YR, Lin HC. Risk of adverse perinatal outcomes with antithyroid treatment during pregnancy: a nationwide population-based study. BJOG: An International Journal of Obstetrics & Gynaecology. 2011;118(11):1365–73.

29. Rosenfeld H, Ornoy A, Shechtman S, Diav-Citrin O. Pregnancy outcome, thyroid dysfunction and fetal goitre after in utero exposure to propylthiouracil: a controlled cohort study. Br J Clin Pharmacol. 2009;68(4):609–17.

30. Hawken C, Sarreau M, Bernardin M, Delcourt AC, Muller A, Lefort G, Pernollet P, Marechaud R. Management of Graves’ disease during pregnancy in the Poitou-Charentes Region. Ann Endocrinol (Paris). 2016 Oct;77(5):570–577.

31. Lo JC, Rivkees SA, Chandra M, Gonzalez JR, Korelitz JJ, Kuzniewicz MW. Gestational thyrotoxicosis, antithyroid drug use and neonatal outcomes within an integrated healthcare delivery system. Thyroid. 2015; 25(6):698–705.

32. Clementi M, Di Gianantonio E, Cassina M, Leoncini E, Botto LD, Mastroiacovo P. Treatment of hyperthyroidism in pregnancy and birth defects. J Clin Endocrinol Metab. 2010; 95(11):E337±41.

33. Lian XL, Bai Y, Xu YH, Dai WX, Guo ZS. Effects of maternal hyperthyroidism and antithyroid drug therapy on congenital malformation of newborn infants. Chin J Obstet Gynecol. 2005; 40: 511–515.

34. Gianantonio ED, Schaefer C, Mastroiacovo PP, Cournot MP, Benedicenti F, Reuvers M. Adverse effects of prenatal methimazole exposure. Teratology. 2001; 64: 262–266.

35. Wing DA, Millar LK, Koonings PP, Montoro MN, Mestman JH. A comparison of propylthiouracil versus methimazole in the treatment of hyperthyroidism in pregnancy. Am J Obstet Gynecol. 1994; 170: 90–95.

36. Momotani N, Ito K, Hamada N, Ban Y, Nishikawa Y, Mimura T. Maternal hyperthyroidism and congenital malformation in the offspring. Clin Endocrinol (Oxf). 1984; 20: 695–700.

37. Genetics Home Reference. 22q11.2 deletion syndrome. Available at: https://ghr.nlm.nih.gov/condition/22q112-deletion-syndrome. Accessed 19/12/2019.

38. Liew Z, Olsen J, Cui X, Ritz B, Arah OA. Bias from conditioning on live birth in pregnancy cohorts: an illustration based on neurodevelopment in children after prenatal exposure to organic pollutants. Int J Epidemiol. 2015 Feb;44(1):345–54.

